# Distinct origins and transmission pathways of *bla*_KPC_ Enterobacterales across three U.S. states

**DOI:** 10.1101/2022.09.15.22279972

**Authors:** Zena Lapp, Rany Octaria, Sean M. O’Malley, Tu Ngoc Nguyen, Hannah Wolford, Ryan Crawford, Christina Moore, Paula Snippes Vagnone, Diane Noel, Nadezhda Duffy, Ali Pirani, Linda S. Thomas, Brittany Pattee, Claire Pearson, Sandra N. Bulens, Sophie Hoffman, Marion Kainer, Melissa Anacker, James Meek, Isaac See, Allison Chan, Ruth Lynfield, Meghan Maloney, Mary K. Hayden, Evan Snitkin, Rachel B. Slayton

## Abstract

**Background:** Carbapenem-resistant Enterobacterales (CRE) are among the most concerning antibiotic resistance threats due to high rates of multidrug resistance, transmissibility in healthcare settings, and high mortality rates. We evaluated the potential for regional genomic surveillance to track *bla*_KPC_-carrying CRE (KPC-CRE) transmission across healthcare facilities in three U.S. states.

**Methods:** Clinical isolates were collected from Connecticut (CT; 2017-2018), Minnesota (MN; 2012-2018), and Tennessee (TN; 2016-2017) through the U.S. Centers for Disease Control and Prevention’s Multi-site Gram-negative Surveillance Initiative and additional surveillance. KPC-CRE isolates were whole-genome sequenced, and case report data on patient comorbidities, healthcare utilization, and interfacility patient transfer were extracted.

**Findings:** In CT, most KPC-CRE isolates showed evidence of importation from outside the state, with limited local transmission. In MN, cases were mainly from sporadic importation and transmission of *bla*_KPC_-carrying *Klebsiella pneumoniae* (KPC-Kp) ST258, and clonal expansion of an imported epidemic lineage of *bla*_KPC_-carrying *Enterobacter hormaechei* (KPC-Ec) ST171 primarily at a single focal facility and its satellite facilities. In TN, KPC-Kp ST258, and more recently emerged KPC-Kp ST307 and KPC-Eh ST114 were most common, with largely non-overlapping facility networks mediating the spread of ST258 versus ST307 and ST114.

**Conclusions:** The underlying processes driving KPC-CRE burden can differ substantially across regions, and different STs can spread via distinct pathways within a region. Integrating genomic and epidemiological data from regional surveillance, and information on interfacility patient transfers, can provide insights to target interventions.

## Introduction

Carbapenem-resistant Enterobacterales (CRE) accounted for an estimated 13,100 infections among hospitalized patients in the United States (U.S.) in 2017 [1], and likely colonized upwards of 100,000 additional patients [2]. Infections with CRE are an urgent public health threat as they can be difficult to treat due to resistance to carbapenems and other last-resort antibiotics [3,4]. Of concern are CRE containing a carbapenemase gene found on a mobile genetic element, which can disseminate within and between different species via horizontal gene transfer (HGT) followed by subsequent clonal dissemination [5]. To implement interventions that effectively reduce CRE infections and transmission requires not only monitoring regional CRE infections, but also understanding where they were acquired. In particular, because of the capacity for CRE to colonize patients for months and even years, the facility where a patient developed an infection may not be where they acquired it. As active surveillance for CRE colonization at a regional level is logistically infeasible, strategies are needed to leverage clinical isolate collections to discern the origin of patient’s CRE infections.

Understanding whether and how a hospitalized patient’s CRE strain is related to previous cases is critical for effective and efficient surveillance and intervention strategies. The CRE burden in some regions appears to be driven by the evolution or importation of lineages with epidemic potential, such as *bla*_KPC_-carrying *Klebsiella pneumoniae* (KPC-Kp) ST258 (e.g. in California) [5] and *Enterobacter hormaechei* (KPC-Eh) ST171 (e.g. in Minnesota) [6] in the U.S. Following initial regional emergence, these strains may be transmitted within healthcare facilities and spread between facilities via transfer of colonized patients [5,7,8]. In regions with sustained transmission of epidemic clones, effective regional control requires identification of locations where transmission is occurring and impacting the burden in the region, and monitoring the movement of CRE carriers between healthcare facilities [9,10]. In contrast, others have observed a role for HGT of mobile carbapenemase elements in driving CRE spread within individual healthcare facilities [11–13]. If HGT is a significant contributor to overall regional CRE burden, effective prevention and control requires identification of reservoirs of these resistance elements and an understanding of the propensity of different strain/mobile element combinations to spread between patients or act as HGT donors.

Here, we evaluate the potential for passive regional genomic surveillance to inform the pathways leading to CRE cases across regional healthcare networks in three U.S. states involved in the U.S. Centers for Disease Control and Prevention’s (CDC) Multi-site Gram-negative Surveillance Initiative (MuGSI). In particular, we focus on the most commonly observed CRE in the MuGSI catchment areas, KPC-Kp and KPC-Ec. This builds off of the MuGSI aim to quantify the burden of certain resistant gram-negative bacteria in the U.S. by supplementing preexisting data with whole-genome sequencing (WGS) of CRE isolates collected through MuGSI and supplemental state-wide surveillance. By integrating genomic data from densely sampled CRE cases and healthcare exposure data, and comparing local strains with global isolates, we gained insight into local strain origins and regional transmission pathways.

## Methods

This retrospective study was a collaboration between the Emerging Infections Program (EIP) and CDC Prevention Epicenters. The EIP is a collaboration between 10 state health departments and their partnering academic institutions and the Centers for Disease Control and Prevention and other federal agencies [14]. The Healthcare-Associated Infections Community Interface (HAIC) is one of the core components of the EIP and conducts active population- and laboratory-based surveillance for CRE [15]. Three EIP sites participated in this study: Connecticut (CT), Minnesota (MN), and Tennessee (TN).

### CRE surveillance

While only selected counties in MN and TN participated in the HAIC MuGSI [16], each of the participating states conducted active statewide public health surveillance for CRE. All carbapenem-resistant isolates were submitted by clinical laboratories to each state’s respective State Public Health Laboratory (SPHL; see Supplemental Methods). Multiple isolates from the same patient were obtained if the new isolate was from at least 30 days after a previous case isolate was identified.

We define CRE as any organism in the Enterobacterales order isolated from any clinical specimen with resistance to doripenem, meropenem, or imipenem (minimum inhibitory concentrations [MIC] of ≥4 μg/ml); with resistance to ertapenem (MIC ≥2 μg/ml); or with demonstrated production of a carbapenemase [16]. All CRE isolates identified were tested for the carbapenemase genes *bla*_KPC,_ *bla*_NDM_, *bla*_OXA-48_, *bla*_VIM_, and *bla*_IMP_ (see Supplemental Methods).

CRE considered for WGS were collected in CT from 2017-2018, in MN from 2012-2018, and in TN from 2016-2017. Unless otherwise noted, only isolates from 2016-2018 were analyzed to enable more meaningful comparison of analyses across states. For CT and MN, isolates from the entire state were included. In TN, only isolates from TN Emergency Medical Services (EMS) Regions 2-7 were included (76.6% of the population). We excluded counties in the Memphis-Delta and Northeast Tennessee Region due to significant healthcare utilization across state lines. **Table S1** provides information on the number of each type of healthcare facility in each state and the number of each type for which we have at least one sequenced isolate.

### Epidemiologic data

Epidemiologic data were obtained through medical record review and in TN supplemented by additional data sources (see Supplemental Methods). Isolate metadata includes state, treatment facility ID, patient ID, age, sex, culture source, and hospitalizations in the previous year. MuGSI surveillance data also included information on infection type and underlying conditions, which was only obtained for selected non-MuGSI isolates. The only time we used this additional data was to investigate shared underlying conditions; this analysis was limited to samples for which we had data on these conditions. All other analyses included all isolates.

### Generation of aggregate patient transfer networks

Aggregate patient transfer networks (i.e., patient flow across healthcare facilities [5]) for each state for 2017 were derived from CMS fee-for-service beneficiary claims data linked to the CMS Minimum Data Set by Medicare beneficiary ID. The number of transfers between two facilities includes transfers directly from one facility to another and transfers with an intervening stay in the community of less than 365 days. See Supplemental Methods.

### Isolate and genomic data processing

Over 90% of clinical CRE isolates considered for WGS were KPC-Kp or KPC-Eh; we therefore performed Illumina WGS on only this subset of isolates (see Supplemental Methods). Study isolates included isolates from BioProject numbers PRJNA272863 [6] and PRJNA873034 (new from this study) (Supplemental File S1). For this set of isolates, we identified species and sequence types (STs) [17,18], called single-nucleotide variants using species-specific reference genomes (*K. pneumoniae*: KPNIH1, GenBank accession number CP008827.1, 5,394,056 base pairs; *E. hormaechei*: MNCRE9, GenBank accession number JZDE00000000.1, 4,911,317 base pairs) [19–25], built reference-based phylogenetic trees [24–28], and generated and annotated assemblies [19,20,29,30] (see Supplemental Methods). These reference-based data were used for all analyses unless otherwise noted. Furthermore, to prevent overcounting of intra-facility transmission, isolates were limited to the first isolate per patient for each ST/facility/patient combination.

### Identification of putative geographic importation events

5,346 public KPC-positive and KPC-negative *K. pneumoniae* and *E. hormaechei* isolates were downloaded from the PATRIC database on 2021-04-23 [18,31] and used to contextualize study isolates. In particular, we aimed to group study isolates into clusters that could be traced back to common local emergence events based on phylogenetic clustering, and then used geographic context to determine whether a cluster likely originated via importation of a *bla*_KPC_ carrying strain from outside the state. For all public and study isolates, ST-specific core genome phylogenies were generated [24,32,33] and *bla*_KPC_ was identified [31,34] (see Supplemental Methods). For each core genome tree, maximum likelihood ancestral reconstruction of each study site location (MN, TN, CT) and presence of *bla*_KPC_ was performed individually in R v4.0.2 [35] using the ace() function in ape v5.5 [36]. Nodes with an ancestral reconstruction confidence of < 0.875 were discarded. All transition events (for each geographic location and *bla*_KPC_) were then mapped onto the edges of the phylogeny. Each KPC-positive study isolate tip was traversed towards the root of the tree until the first transition event was identified. If the first transition event was a geographic transition event with no *bla*_KPC_ transition event on the same edge, isolates were considered to originate from geographic importation of a KPC-positive strain. If the first transition event was for *bla*_KPC_, this supports local acquisition of *bla*_KPC_ via HGT; therefore, these isolates were presumed to not originate from importation of a KPC-positive strain. If a geographic and *bla*_KPC_ transition occurred on the same edge, or if transition edges were of low confidence, we considered there to be no evidence of importation of a KPC-positive strain. Study isolates traced back to the same edge transition event, regardless of the type or confidence of the transition, were considered part of a local cluster traced back to a common local emergence event. We consider the number of transition events for a given geographic location to be the number of identified clusters.

### Computing pairwise distances

Pairwise single nucleotide variant (SNV) distances and patristic distances for study isolates from each species-specific reference-based sequence alignment were calculated in R v4.0.2 [35] using the dist.dna() function in ape v5.5 [36] with pairwise.deletion = TRUE. These pairwise SNV distances were classified as intra- and inter-facility pairs using the get_pair_types() function in regentrans v0.1 [37]. Rather than choosing a specific pairwise SNV distance threshold, we highlight various thresholds (≤ 5, 10, 15) as a sensitivity analysis. A threshold of 10 was chosen where needed based on a decrease in the fraction of intra-facility pairs at various SNV distances (**Figure S3A**).

### Integrated genomic and epidemiologic analyses

#### Relatedness of intra- vs. inter-facility isolates

For each isolate with at least one intra-facility isolate pair and at least one inter-facility isolate pair, we identified the minimum pairwise SNV distance of all intra-facility pairs and the pairwise minimum pairwise SNV distance of all inter-facility isolate pairs. We then compared the pairwise SNV distances of intra- vs. inter-facility isolate pairs using a paired Wilcox test.

#### Intra-facility isolate pair shared comorbidity analysis

For each intra-facility isolate pair, we determined whether they had at least one shared comorbidity and used a Wilcox test to compare pairwise SNV distances of patients who shared comorbidities and those who did not. This was done based on the assumption that patients with shared comorbidities would be provided care in more proximate inpatient locations.

#### Inter-facility isolate pair shared prior healthcare exposure analysis

For each inter-facility patient pair, we determined whether they had a shared healthcare exposure in the past year (data collection method described in the Supplemental Methods). Then we used a Wilcox test compared the pairwise SNV distances of patients who shared a healthcare exposure and those who did not.

### Data analysis and visualization

R v4.1.1 was used for all data analysis and visualization using these packages: tidyverse v1.3.1 [39], ggtree v2.5.2 [40,41], igraph v1.2.6 [42], regentrans v0.1 [37], tidygraph v1.2.0 [43], ggraph v2.0.5 [44], exact2×2 v1.6.5 [38], cowplot v1.1.1 [45], readxl v1.3.1 [46], ape v5.5 [36]. The code used to perform analyses is here: https://github.com/Snitkin-Lab-Umich/eip-cre-transmission-ms.

### Human subjects research

The study was reviewed and approved by the Tennessee Department of Health institutional review board (IRB). See 45 CFR. part 46; 21 CFR part 56. The Minnesota and Connecticut Department of Health IRB determined that the study was exempt from IRB review in accordance with 45 CFR 46.101(b)(4). The study was approved by the CDC IRB with a waiver of HIPAA authorization under the Privacy Rule as per 45 CFR 46.512(i). The University of Michigan Medical School IRB approved this protocol.

## Results

Over 90% of collected clinical CRE isolates were KPC-Kp or KPC-Eh; we therefore focus our analysis on this subset of isolates to understand the pathways leading to the dominant causes of infections. We included the first isolate of each unique ST/facility/patient combination. In each state, over 50 KPC-Kp and KPC-Eh isolates were present across over 20 distinct healthcare facilities (**Table S1**; **Figure S1**). The dominant STs were KPC-Kp ST258 in all three states, KPC-Eh ST171 in MN, and KPC-Eh ST114 and KPC-Kp ST307 in TN.

### Clonal dissemination after importation or *bla*_KPC_ acquisition differs across states and STs

To gain insight into the origin and magnitude of spread of different KPC-Kp and KPC-Eh strains in each state we performed phylogenetic analyses including 5,346 publicly available *Enterobacter* spp. and *Klebsiella* spp. genomes deposited in the PATRIC database (**Figures S2A**-**B**). We sought to use the genetic and geographic context provided by PATRIC isolates to identify phylogenetic clusters including only isolates from a given state (**Figure 1**; **Figure S2C**). Despite what we assume to be relatively sparse sampling of circulating *Klebsiella* and *Enterobacter* strains in PATRIC, phylogenetic context revealed that KPC-Kp and KPC-Eh cases in all three states were attributable to large numbers of independent regional emergence events. In CT and MN, large phylogenetic clusters consistent with local dissemination were almost exclusively restricted to those that could be traced back to the outside importation of the epidemic lineages KPC-Kp ST258 and KPC-Eh ST171. In TN, we also detected evidence for local dissemination of imported KPC-Kp ST258, and to a lesser extent KPC-Eh ST171. However, in contrast to CT and MN, we also found large phylogenetic clusters that could not be traced to an external source. Most notable were large clusters of the emerging epidemic clones KPC-Kp ST307 [48] and KPC-Eh ST114 [49]. Support for these phylogenetic clusters being due to local acquisition of *bla*_KPC_ comes from the nearest neighbors of these strains in public collections being KPC-negative (**Figure S2C**).

**Figure 1:**
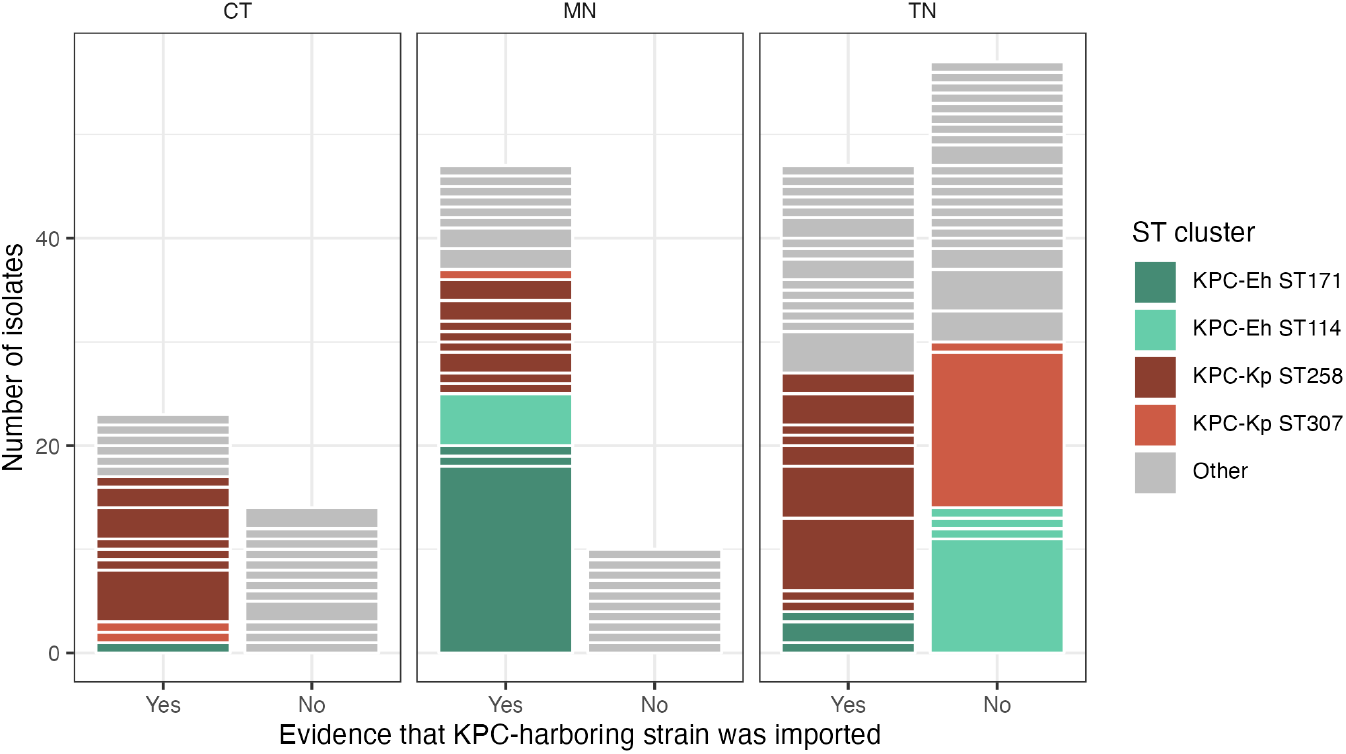
Evidence of importation and clonal dissemination differs across states and sequence types (STs). Clusters are separated by white lines between the colored bars, where the bar height of a given cluster is the cluster size. Clusters were defined as subclades that were monophyletic for a single state. Importation events were considered those with a KPC+ edge transition from another state to the EIP state (see Methods). We observed events with limited onward transmission in all states. We also see a large cluster of *E. hormaechei* ST171 in MN that arose from a putative importation event, indicating importation followed by sustained dissemination. Additionally, we observed large clusters of *E. hormaechei* ST114 and *K. pneumoniae* ST307 in TN with no evidence of importation from another state. CT=Connecticut; MN=Minnesota; TN=Tennessee; ST=sequence type; Eh=*E. hormaechei*; Kp=*K. pneumoniae*.

### Genomic analysis of regional clinical isolate collections allowed for the detection of local transmission in MN and TN, and lack thereof in CT

Having characterized the origin of circulating strains, we next sought to understand whether regional genomic surveillance of clinical isolates was dense enough to discern facilities and regional subnetworks where transmission was occurring. To this end, we first investigated the extent to which the surveillance isolates captured putative recent local transmission events, defined as small pairwise SNV distances (**Figures S3B**) or small patristic distances (**Figure S3C**). Within an ST, smaller pairwise SNV distances indicate potential local transmission, while larger distances are not indicative of local transmission [50]. We captured very few local transmission events in CT, suggesting that importation from other geographic regions may be driving the case load there. In contrast, we captured likely local transmission events of KPC-Eh ST171 in MN and of KPC-Kp ST258, KPC-Kp ST307 and KPC-Eh ST114 in TN, consistent with our observation of large phylogenetic clusters for these STs (**Figure 1**).

Patients with closely related intra-facility isolate pairs did not share any underlying conditions that may be indicative of an outbreak related to a certain location or procedure in the hospital (all p > 0.05). In addition, we observed that isolates close in pairwise SNV distance to another isolate from the same facility were also often closely related to an isolate from a different facility (**Figure 2A)**. In fact, for ST258 and ST114 in TN, isolates tended to have closer genetic neighbors at different facilities versus the same facility (p < 0.05). These findings suggest that observation of closely related isolates from within a facility does not directly equate to an intra-facility transmission event having occurred, but may also reflect recent transmission at a connected facility. Supporting the notion that we observed signatures of inter-facility transmission is that patients from different facilities with a common prior healthcare exposure tended to have more closely related isolates than those with no common prior healthcare exposure (**Figure 2B**).

**Figure 2:**
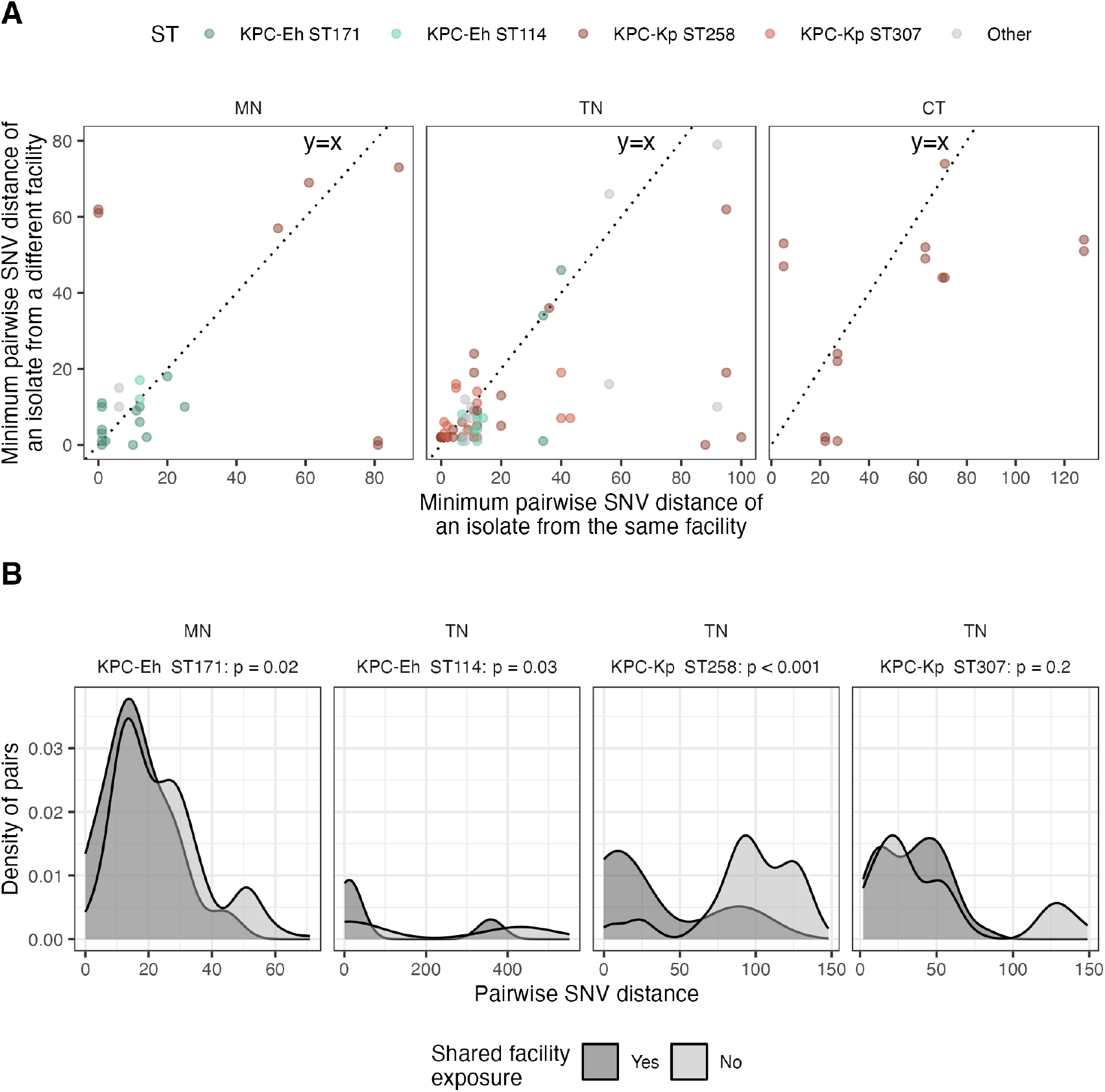
Small pairwise single nucleotide variant (SNV) distances between isolates suggest that local transmission was captured. (A) For each isolate with at least one intra-facility isolate pair and at least one inter-facility isolate pair, pairwise SNV distance of the most closely related intra-facility compared to the most closely related inter-facility isolate. (B) Pairwise SNV distance of inter-facility isolate pairs from patients with a shared facility exposure compared to those not linked by a shared facility exposure. A shared facility exposure is when individuals with isolates from different facilities both spent time in the same facility at some point in the last year. Only STs with >10 isolate pairs with a pairwise SNV distance of ≤15 are shown. One-sided Wilcox p-values compare pairwise SNV distances of isolates with a shared facility exposure (N ranges from 14 to 35) to isolates without a shared facility exposure (N ranges from 67 to 376). CT=Connecticut; MN=Minnesota; TN=Tennessee; SNV=single nucleotide variant; ST=sequence type; Eh=*E. hormaechei*; Kp=*K. pneumoniae*.

### Longitudinal genomic surveillance in MN revealed limited spread of ST258 and localized transmission of ST171 in a regional sub-network

With evidence for isolate pairs with small SNV distances being informative of recent intra- and inter-facility transmission, we next set out to look at genomic linkages across MN and TN to gain insight into where transmission of different STs is occurring. MN has been collecting CRE isolates since 2012. We observed a decrease in the amount of KPC-Eh ST171 and KPC-Kp ST258 (**Figure S4A**) and corresponding signatures of transmission over time (**Figure S4B**). Incorporating information from this extended dataset to investigate transmission, we observed stark contrasts in the relatedness of ST171 compared to ST258 isolates across time. ST258 isolates collected over one year apart were rarely closely related to each other; however, ST171 isolates collected even over 5 years apart were often quite closely related (**Figure 3A**). We investigated whether this difference was driven by certain facilities, and found that one facility, F38, was highly represented in closely related ST171 intra- (**Figure 3B**) and inter-facility (**Figure 3C**) isolate pairs. Of all ST171 isolate pairs within 10 SNVs, 66/86 (77%) intra-facility pairs were from F38 and 240/430 (56%) inter-facility pairs contained at least one isolate from F38. About half of these inter-facility pairs were connected to one of three other facilities (**Figure 3D**). F38 is highly connected to other facilities in the patient transfer network (**Figure S3E**) and is well-connected by patient transfer to the other three highly represented facilities (**Figure S3F**; ≥90^th^ percentile of all facility pairs). These findings indicate that the pattern by which ST171 and ST258 disseminated, and the potential role of specific facilities, is very different.

**Figure 3:**
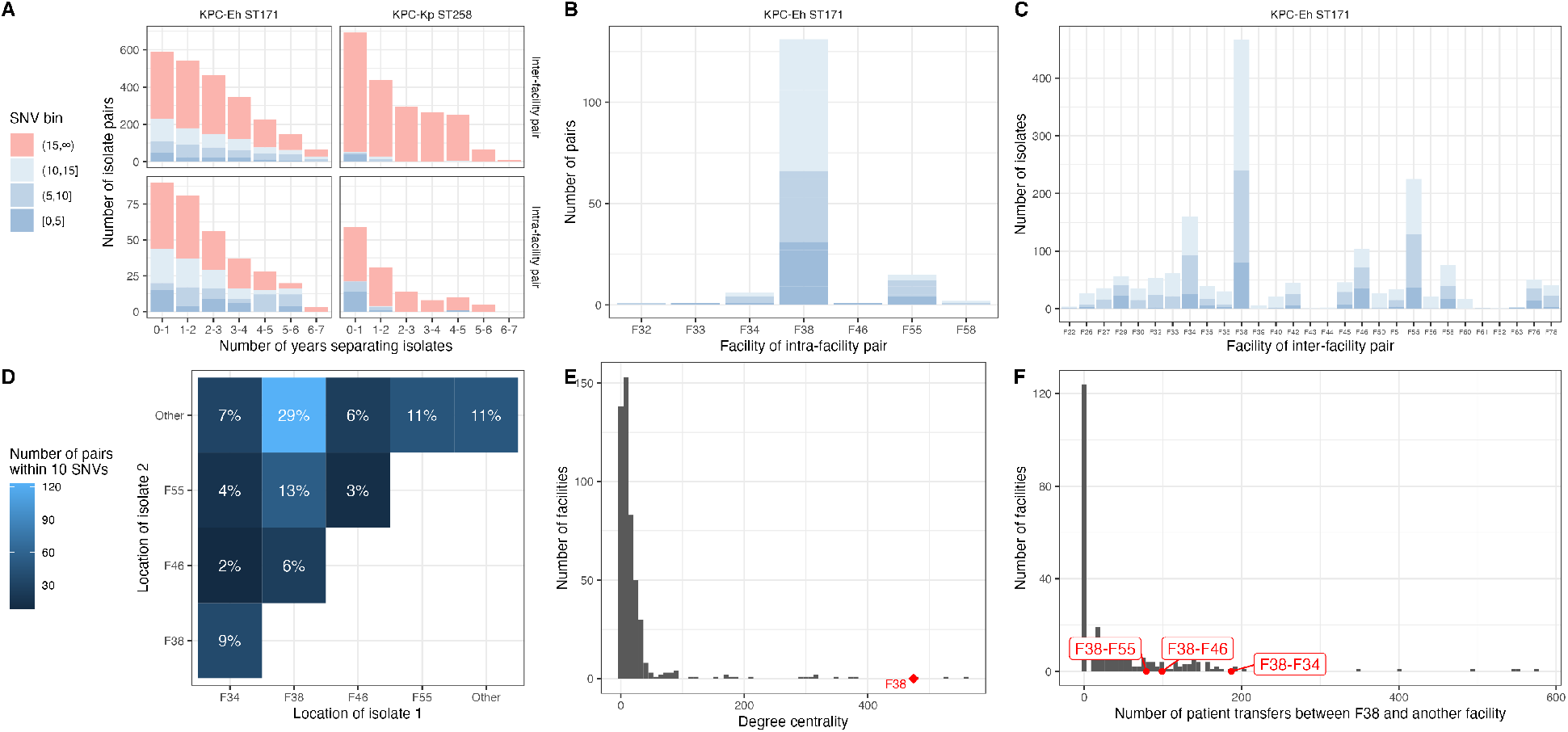
Transmission of KPC-Eh ST171 in Minnesota (MN). (A) Number of years separating ST171 and ST258 isolate pairs. (B) Intra-facility ST171 isolate pairs by facility. Each pair is represented once. (C) Inter-facility isolate pairs by facility. Each pair is represented twice (once per facility). (D) Facilities connected by ST171 isolates within 10 SNVs of each other. (E) Degree centrality of all facilities, with F38 highlighted. The in- and out- degree values were summed (from and to a facility). (F) Number of patient transfers between F38 and each other facility. The values to and from each facility were summed. SNV=single nucleotide variant; ST=sequence type; Eh=*E. hormaechei*; Kp=*K. pneumoniae*.

### Distinct facility sub-networks in TN harbor different lineages

We next explored whether we could discern where transmission was occurring in TN by investigating what facilities patients with each ST had been to in the previous year. We observed a cluster of seven facilities where patients with ST114, ST307, and as STs categorized as Other, had been in the previous year (**Figure 4A**). Visualizing these seven facilities in the context of the aggregate patient transfer network revealed that they are closely connected (**Figure 4B**) and have more patient sharing among themselves than with other facilities (**Figure 4C**; Wilcox p < 0.001). Additionally, this facility cluster is distinct from a cluster containing predominately ST258 (**Figure 4B**).

**Figure 4:**
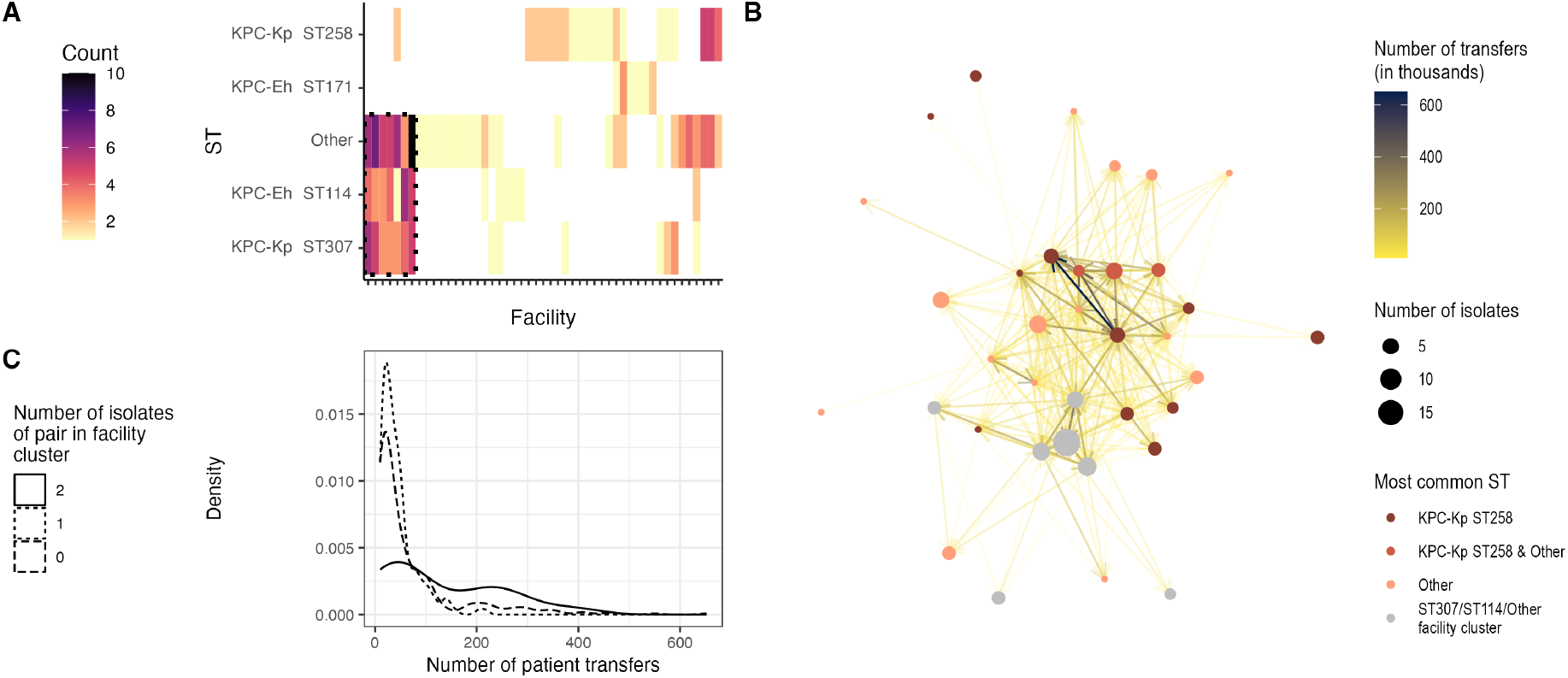
Distinct facility sub-networks in Tennessee (TN) harbor different lineages. (A) Number of patient facility exposures in the prior year for each facility and ST. Each patient may be represented more than once if they have more than one facility exposure. A cluster of facilities identified using the complete linkage clustering method (dashed box) are common in patients with ST114, ST307, and other isolates. (B) Patient transfer network of facilities with at least one whole-genome sequenced isolate plotted using the Kamada-Kawai algorithm. The cluster of facilities identified in the heatmap in panel A (Kp ST307/Ec ST114/Other facility cluster) are also clustered in the network. (C) Number of patient transfers for facility pairs where 0, 1, or 2 of the facilities are in the facility cluster identified in the heatmap in panel A. Facility pairs where both facilities are in the cluster have more patient sharing than facility pairs where only 0 or 1 of the facilities is in the identified cluster. ST=sequence type; Eh=*E. hormaechei*; Kp=*K. pneumoniae*.

## Discussion

As public health laboratories increase their capacity for genomic surveillance, it is critical to understand if and how genomic data from passive surveillance can be used to guide regional intervention efforts. Statewide active surveillance is infeasible, and while that would be required to infer who transmitted to whom, passive surveillance may be enough to provide practical guidance for interventions including if and where transmission is happening and if there are any emerging threats. Here, we evaluated whether genomic analysis of clinical *bla*_KPC_-positive CRE isolates from three U.S. states could inform our understanding of the origins and transmission pathways of circulating strains. We could discern the relative contributions of importation and clonal dissemination to each state’s KPC-Kp and KPC-Eh case burden and highlight individual facilities and connected regional sub-networks where putative transmission occurred. These insights did not rely on detailed clinical metadata, indicating that other regions interested in identifying hotspots and pathways for regional spread may be able to do so with only knowledge of recent healthcare exposures. Our findings suggest that states should consider prioritizing genomic analysis of clinical isolates to monitor transmission hotspots as insights gleaned from these analyses may enable targeting of regional infection prevention efforts to certain facilities.

Despite focusing on the same two Enterobacterales species harboring the same carbapenemase gene, we observed stark differences in the underlying drivers of each state’s KPC-Kp and KPC-Eh burden. In CT, strains were frequently imported but did not show evidence of onward transmission. Importation is unsurprising, given the state’s proximity to other major CRE hotspots in the Northeast U.S. [51]. However, this still leaves unanswered why imported *bla*_KPC_-positive CRE have apparently not gained a foothold. One reason may be that the structure of the CT regional healthcare network decreases local transmission. Recent work showed that post-acute care settings can act as initiators and amplifiers of regional CRE epidemics [52,53]. Thus, infection prevention practices in these facilities, and their patient transfer connections with other facilities, may have large impacts on overall regional CRE spread. Regardless, our observations suggest that focusing resources on testing admission cultures from out-of-state may allow CT to continue having a low burden of these CRE strains.

In MN, *bla*_KPC_-positive CRE burden was driven by a major importation event of the epidemic lineage KPC-Eh ST171 followed by dissemination within the region. Incorporation of historical data revealed that isolates far apart in time were often closely related and were largely from or connected to isolates from a single hub facility with many referrals that provides specialty care, F38. These observations could indicate an environmental reservoir within or proximate to F38, or undetected endemic spread at one or more referral facilities. Having this data in real-time generates actionable hypotheses to potentially stem regional spread. F38 could prioritize testing patients admitted from facilities connected by extensive patient transfer. Similarly, state epidemiologists could investigate facilities with large numbers of genomic linkages to isolates from F38, such as the three identified here, to detect other potential reservoirs and locations where intervention may have a broad positive impact on regional KPC-Eh burden.

TN showed a diversity in circulating strains. In addition to a significant burden of ST258, we observed CRE isolates from two more recently emerged lineages – KPC-Kp ST307 and KPC-Eh ST114 – and less-common STs that appear to have emerged and spread more recently. This pattern is in stark contrast with CT and MN, where there were very few cases of onward transmission detected after a non-epidemic strain acquired *bla*_KPC_. Furthermore, these recently emerged lineages traversed a distinct sub-network of healthcare facilities relative to KPC-Kp ST258. These observations suggest that the propensity for potential HGT of *bla*_KPC_ into emerging and novel receptive strains, followed by clonal dissemination, appears to be higher in a subset of TN healthcare facilities. While we were unable to discern specific HGT events in this study, nor why these facilities may be hotspots for successful HGT, our observations do suggest that the subnetwork observed here may be an important location to monitor for emerging threats.

Our study has several limitations. First, our use of only clinical isolates may lead to an underestimation of clonal dissemination, that may be different for different STs depending on their virulence and associated colonization-to-infection ratio (i.e. ST-specific iceberg effect). However, we were still able to uncover patterns of clonal dissemination using these data, suggesting that regional clinical isolate collections can provide actionable insights for public health labs. While transmission may not have occurred at the facilities where clinical isolates were collected, this nevertheless provides a location at which to initiate an investigation and could ultimately guide more targeted surveillance efforts. Second, the isolate collection periods were different for each state. This may result in different levels of contextual information provided by public databases, through which importation was inferred. However, our observations are consistent with prior studies in that KPC is known to be stably associated with ST258, ST171 emerged in the upper Midwest, and ST307 and ST114 only recently became associated with KPC. Another limitation is that the public isolates used from PATRIC to inform the origin of locally circulating strains are not equally representative of all geographic regions, which may underestimate the extent of importation, as multiple importations from an unsampled reservoir may be merged. While the analyses performed here would be enhanced by additional context, we note that inferences made into importation using phylogenetic clustering are largely consistent with the SNV distance-based analyses, supporting the robustness of our conclusions. Furthermore, the aggregate patient transfer data used to understand the connectivity between regional healthcare facilities was derived from Medicare patient transfers, which may lead to biases in patient transfer connections. However, despite this, we were able to discern local sub-networks of healthcare facilities with distinct transmission patterns.

In conclusion, our results suggest that, not only can the origins and transmission patterns of *bla*_KPC_-positive CRE be investigated using genomic surveillance of sufficiently comprehensive regional clinical isolate collections, but also that these transmission dynamics can vary across strains and regions. The differences underlying the KPC-Kp and KPC-Eh burden in these three states would not have been discernible without genomic data, and in fact would be strengthened further by the existence of larger genomic surveillance initiatives that could provide more granular context for the origin of strains and their mobile elements to guide prevention efforts. Similarly, our understanding of how KPC-Kp and KPC-Eh proliferate in a region would be limited without knowledge of patient healthcare exposures and transfer networks, which enable a more nuanced understanding of where transmission is occurring, where reservoirs might exist, and where additional surveillance or intervention is required. Taken together, these observations support the value of robust regional genomic surveillance for antibiotic resistance threats and the need for analytic platforms capable of integrating genomic and patient movement data to guide local and state infection prevention efforts.

## Data Availability

Isolates from BioProject numbers PRJNA272863 [6] and PRJNA873034 (new from this study) were used.
The code and shareable data used to perform all analyses can be found here: https://github.com/Snitkin-Lab-Umich/eip-cre-transmission-ms.

https://www.ncbi.nlm.nih.gov/bioproject/PRJNA272863

https://www.ncbi.nlm.nih.gov/bioproject/PRJNA873034

https://github.com/Snitkin-Lab-Umich/eip-cre-transmission-ms

## Acknowledgements

Pamela Talley and Matt Estes from the Tennessee Department of Health. The MN-PHL Microbiology and Sequencing & Bioinformatics laboratories for their work in identifying and sequencing all of the MN isolates in the project. Gwendolyn Hughes and Christina Sancken from CDC.

## Disclaimer

The findings and conclusions in this report are those of the authors and do not necessarily represent the views of the Centers for Disease Control and Prevention. The use of trade names is for identification only and does not imply endorsement by the Centers for Disease Control and Prevention.

## Supplemental information

### Supplemental methods

#### Catchment areas

The Multi-site Gram-negative Surveillance Initiative (MuGSI) [16] CRE surveillance catchment areas for each state participating in this study are: the entire state of CT; Hennepin and Ramsey counties, encompassing the metropolitan Minneapolis-St. Paul area, of MN; and TN EMS Region 5, an 8-county region encompassing the metropolitan Nashville area, of TN. The TN the Memphis-Delta and Northeast Regions were excluded due to healthcare utilization across state lines.

#### CRE surveillance

CRE are defined as any organism in the Enterobacterales order isolated from any clinical specimen and resistant to doripenem, meropenem, or imipenem (minimum inhibitory concentrations [MIC] of ≥4 μg/ml), ertapenem (MIC ≥2 μg/ml), or demonstrates production of a carbapenemase [16]. Specifically, isolates from residents of the surveillance area identified at the clinical laboratory as CRE from *Escherichia coli, Enterobacter cloacae* complex species (i.e., *E. cloacae, E. asburiae, E. bugandensis, E. hormaechei, E. kobei, E. ludwigii*, and *E. nimipressuralis*), and *Klebsiella* species (i.e., *K. aerogenes, K. oxytoca*, and *K. pneumoniae*) were included. All CRE isolates identified through the described public health surveillance systems were characterized at the respective SPHL, which tested for carbapenemase genes (e.g., *bla*_KPC,_ *bla*_NDM_, *bla*_OXA-48_, *bla*_VIM_, *bla*_IMP_; see below for more details). While all CRE are supposed to be tested for carbapenemases, this does not always happen in practice; the percentage of CRE tested for carbapenemases was 81.5% in CT, 90% in MN, and 78% in TN.

##### Connecticut

CT did not participate in MuGSI surveillance during the study period, so all study isolates from CT came from statewide passive surveillance for CRE. CRE surveillance of all Enterobacterales isolates collected from all invasive clinical sites, respiratory sources, and urine began on Jan 1, 2017. All CRE isolates undergo antimicrobial susceptibility testing with a custom Sensititre and disk diffusion panel, phenotypic detection of carbapenemase activity using the mCIM test and PCR detection of carbapenemase genes (*bla*_KPC_, *bla*_NDM_, *bla*_OXA-48_, *bla*_VIM_, *bla*_IMP_). Isolates carrying *bla*_KPC_ that were identified in 2017 and 2018 from this collection were included.

##### Minnesota

MuGSI surveillance in Minnesota is conducted in Ramsey and Hennepin counties. MuGSI isolates are obtained from sterile sites or urine and include *E. coli, Klebsiella* spp., and *Enterobacter* spp. Non-MuGSI isolates are collected from a variety of culture sources (e.g., blood, urine, respiratory) and represent multiple Enterobacterales species. Isolate identification is confirmed by the MALDI Biotyper CA System (Bruker Daltonics, Inc., Billerica, MA). All CRE isolates are tested for carbapenemase production using the modified carbapenem inactivation method (mCIM). Isolates demonstrating phenotypic carbapenemase production undergo molecular testing (i.e., polymerase chain reaction [PCR]) for detection of carbapenemase genes *bla*_KPC_, *bla*_NDM_, *bla*_OXA-48_, *bla*_VIM_, *bla*_IMP_. All *bla*_KPC_ -positive *K. pneumoniae* and *E. cloacae* complex isolates submitted to MN PHL from 2012-2018 were analyzed for this study.

##### Tennessee

EMS region 2-7 were selected because it provides a good representation of patient referrals; captures most isolates in patient transfer networks covering the area; and includes both the Knoxville area in which CRE emerged in 2016, and the Nashville area (the MuGSI catchment area: Dickson, Cheatham, Robertson, Sumner, Wilson, Rutherford, Williamson and Davidson counties) where CRE increased in 2017. CRE isolates underwent organism confirmation using the MALDI-TOF bioMerieux. Isolates were tested for antimicrobial resistance with the Kirby Bauer method. Those isolates that were resistant to at least one carbapenem were then tested by the CarbaNP Method for carbapenemase production. Those that demonstrated carbapenemase production from the CarbaNP underwent a PCR assay developed by the CDC to test for the *bla*_KPC_ and *bla*_NDM_. All *bla*_KPC_-positive isolates collected during 2016–2017 and submitted to the TN SPHL were included.

#### Epidemiologic data

MuGSI surveillance data were collected through medical record review by EIP surveillance officers in each site. Additional details on data collection for isolates identified outside of the MuGSI program are listed below for each site.

##### Connecticut

Clinical case details were abstracted from medical records by EIP surveillance officers with the Connecticut Department of Public Health.

##### Minnesota

At the time of culture collection, surveillance staff completed CRF through electronic or in-person medical chart review.

##### Tennessee

For all carbapenemase-producing Enterobacterales cases, a questionnaire is completed by the healthcare facility of origin. For cases that did not have a completed questionnaire, hospitalization details were gathered from the Statewide Hospital Discharge data (described below).

#### Facility identification

Facility identification was performed through chart review at the reporting facility. State-specific details are described below.

##### Connecticut

Chart review was performed at the reporting facility for all inpatient and hospital network events. When additional referring facilities (including nursing homes, long term acute care hospitals, etc.) were identified on medical record review, follow-up investigation was performed at the additional facilities. Upon identification of outpatient isolates, patient details were collected from ordering providers by phone call and/or faxed data collection instrument.

##### Minnesota

MN staff maintain a compendium with facility name and type according to CMS. The facility name and type are recorded in the MN surveillance database for each isolate and verified using this compendium. For MuGSI cases, chart review at the facility where the isolate was collected was also performed by MN staff.

##### Tennessee

TN staff linked patient identifiers of each case in the National Electronic Disease Surveillance System (NEDDS)-Base System (NBS) surveillance data to the TN Hospital Discharge Data System (HDDS) to identify and confirm the healthcare facility where they had the first specimen culture collected for each CP-CRE case in TN. Alternatively, if the patient was not hospitalized in a TDH-licensed hospital and therefore, was not found in the HDDS, TN staff used surveillance data from NBS and the afore-mentioned questionnaire to identify the healthcare facility of origin. Using this approach, the healthcare facility of origin was identified for 139 (89%) of the 157 included isolates. TN staff also gathered information on healthcare exposures within one year prior to specimen collection by linking the patient identifiers in the surveillance data with the inpatient and outpatient HDDS dataset. Isolates with no healthcare exposure information were not used for the patient sharing network analysis.

#### Linkage/matching of facility names to CMS ID

Linkage of facility names to CMS ID was performed at each respective SPHL.

##### Connecticut

Facilities were matched with CMS IDs using CMS Provider of Services Files available at: https://www.cms.gov/Research-Statistics-Data-and-Systems/Downloadable-Public-Use-Files/Provider-of-Services

##### Minnesota

In MN, CMS IDs were manually entered for each isolate using Excel.

##### Tennessee

In TN, the name healthcare facility of origin was linked with the registered facility name by CMS ID using the SPEDIS and COMPGED commands in SAS 9.4 (SAS Institute, Cary, North Carolina) to allow for inexact matches that accounted for minor spelling variations between the facility names from TN’s surveillance data and those from the CMS dataset. Due to the data-sharing agreement between TDH and the University of Michigan, the facility names and CMS IDs in the WGS dataset and patient transfer data from CMS were de-identified before sharing with The University of Michigan.

#### Generation of aggregate patient transfer networks

Aggregate patient transfer networks for each state for the year 2017 were derived from Centers for Medicare and Medicaid Services (CMS) fee for service beneficiary claims data linked to the CMS Minimum Data Set (MDS) by Medicare beneficiary ID. The dataset comprises a network of medical facilities, including but not limited to short-term acute care hospitals, skilled nursing facilities and long-term acute care hospitals, but may not be representative of non-Medicare beneficiaries. To create an annual transfer network, CDC staff linked each unique beneficiary’s healthcare utilization. This included CMS inpatient claims for acute care stays and the MDS to ascertain a beneficiary’s presence in a nursing home, regardless of payer for that stay. Transfer networks were comprised of the year of interest and looked back to the prior year to obtain all discharging facilities where patients had an opportunity to acquire CRE.

Transfers are defined as a patient discharged from a facility and then subsequently admitted to a facility. The number of transfers between two facilities includes transfers directly from one facility to another as well as transfers with an intervening stay in the community of less than 365 days. We chose to use a 365-day time interval rather than a smaller time interval because this gives us more resolution in comparing the extent of patient flow between facilities. Any facility pairs with 10 or fewer patient transfers were censored, as per data use agreements between CDC and CMS.

For each state, we subset the aggregate patient transfer network to only include the facilities for which we have at least one WGS isolate or ones that are connected to one of these facilities by at least one patient transfer event over the course of a year. We then identified paths of maximum patient flow between this subset of facilities using the get_patient_flow() function in regentrans v0.1 [37].

#### DNA extraction

##### Connecticut

Total DNA extractions performed by CT DPHL for WGS were either by the manual method or automated on the QIAcube. Manual extractions from pure single colony Gram-negative bacterial isolates by QIAgen DNeasy Blood and Tissue kit and follow-up quality control of all purified extracts were conducted following PulseNet Protocols for Whole Genome Sequencing on the Illumina MiSeq system. Automated extractions of bacterial cell pellets were performed using the QIAcube protocol for Isolation of DNA from Gram-negative bacteria and QIAGEN DNeasy Blood and Tissue kit. To obtain the cell pellets, colonies from overnight plates were resuspended in 1x Phosphate buffered saline (PBS, Sigma) to an OD600 of 0.35-0.40. Two milliliter cell suspension were centrifuged at 5,000 *x g* for 5 minutes at 2-8 °C and used for extractions.

##### Minnesota

DNA extraction of the isolates was performed in MN using either the QIAcube (Qiagen, Germantown, MD) extractor and QIAmp DNA Mini extraction kit (Qiagen, Germantown, MD) or the Roche MagnaPure LC and the TNA Isolation extraction kit.

##### Tennessee

DNA extraction of the isolates was performed in TN on a Thermo Fisher, KingFisher Flex auto-extraction instrument, using an Applied Biosystems MagMAX DNA Multi-Sample Ultra extraction kit.

#### Whole-genome sequencing (WGS)

The extracted DNA was prepared for WGS using Illumina Nextera XT library Prep kits with Illumina, Nextera XT Index kits (MN, TN, CT) or with Illumina DNA Prep kit and Illumina CD Indexes (CT) (Illumina, San Diego, CA). The Library preparation process was performed on a Beckman Coulter Biomek Fxp instrument (Beckman Coulter, Brea, CA) (TN), on the Eppendorf epMotion (CT) (Eppendorf, Enfield, CT), or manually (MN, CT). The prepared libraries were sequenced on an Illumina MiSeq sequencing instrument (Illumina, San Diego, CA) using 2×250 (MN, TN, CT) or 2×150 (CT) paired end sequencing with V2 chemistry. The fastq files generated were transferred to the University of Michigan for processing and analysis.

#### Genomic data processing

All genomic data processing and analysis was performed by members of Evan Snitkin’s lab at the University of Michigan.

##### Species/ST identification

MLST was assigned with Ariba v2.14.4 [17]. For PATRIC isolates mlst were extracted from the isolate metadata [18].

##### Sequence alignments

Single-nucleotide variants were called in study sequences using SNPKIT (https://github.com/Snitkin-Lab-Umich/snpkit). Quality of reads was assessed with FastQC v0.11.9 [19]. Adapter sequences and low-quality bases were removed with Trimmomatic v0.36 [20]. Variants were identified by mapping filtered reads to the KPNIH1 reference genome (GenBank accession number CP008827.1; 5,394,056 base pairs) for *K. pneumoniae* sequences and the MNCRE9 reference genome (GenBank accession number JZDE00000000.1; 4,911,317 base pairs) for *E. hormaechei* sequences using bwa v0.7.17 [21], removing polymerase chain reaction duplicates with Picard 2.21.7 [22], removing clipped alignments using Samclip 0.4.0, and calling variants with SAMtools v1.11and bcftools [23]. Variants were filtered from raw results using GATK’s VariantFiltration v3.8 (QUAL, >100; MQ, >50; ≥10 reads supporting variant; and FQ, <0.025) [25]. Consensus files generated during variant calling were recombination filtered using Gubbins v3.0.0 [24]. The alleles at each position that passed filtering were concatenated to generate a non-core variant alignment relative to the respective reference genome. Alleles that did not pass filtering were considered unknown (denoted as N in the alignment).

##### Reference-based phylogenies

A custom Python script was used to filter out (mask) single nucleotide variants in the whole-genome alignment that were: (i) <5 base pairs (bp) in proximity to indels that were identified by GATK HaplotypeCaller [25], (ii) in a recombinant region identified by Gubbins v3.0.0 [24], (iii) in a phage region identified by the Phaster web tool [27] or (iv) they resided in tandem repeats of length greater than 20bp as determined using the exact-tandem program in MUMmer v3.23 [26]. This whole-genome masked variant alignment was used to reconstruct a maximum likelihood phylogeny with IQ-TREE v1.6.12 using the general time reversible model GTR+G and ultrafast bootstrap with 1000 replicates (-bb 1000) [33].

##### Genome assemblies

Quality of reads was assessed with FastQC v0.11.9 [19]. Adapter sequences and low-quality bases were removed with Trimmomatic v0.36 [20]. The clean reads were assembled with Spades v3.14.1 (careful mode) [29]followed by Pilon v1.23 assembly correction [30]. Contigs smaller than 500bp were discarded.

##### Public isolates

All assemblies from the species identified within the EIP dataset were downloaded from the PATRIC database on 04/23/2021 [18]. These data were merged with assemblies from PRJNA603790, PRJNA690239, PRJNA401340, and PRJNA415194 for a total of 74,367 assemblies. Genome annotations for the dataset were generated with RAST v1.035 [31].

##### Core genome phylogenies

A concatenated gene alignment of core genes was generated for the entire dataset with cognac v1.0 [32]. Relevant isolates for understanding the population structure and geographic distribution of the isolates collected as part of the EIP study were identified as any isolate within 25 substitutions or the nearest phylogenetic neighbor by alignment distance. This yielded a subset of the 5,346 isolates, including study isolates and public isolates; and a second, concatenated, core gene alignment was generated with cognac for these most relevant isolates, returning the nucleotide alignment for more precise distance comparisons. To generate sequence type specific alignments, these data were divided into relevant clonal groups. Any clonal group with less than 10 members was merged into a group of its respective species. Outgroups were identified as the isolate with the lowest substitution distance to an isolate assigned to that clonal group or species, for clonal group or species alignments, respectively. A core, concatenated gene alignment for each set of isolates corresponding to species/clonal group was generated using cognac with the corresponding outgroup specified, returning the nucleotide alignment.

Recombinant positions were removed with Gubbins v3.0.0 [24]. For alignments with more than 1000 isolates included, the alignments were randomly downsampled into subsets of 250 sequences, including the outgroup in each alignment, and gubbins was run on the downsampled alignment. This was performed in triplicate, and any position identified as recombinant in the three runs was masked from the alignment. The recombination filtered alignments were then input to IQ-TREE v1.6.12 [33] to generate maximum likelihood phylogenies.

##### bla_KPC_ identification

The *bla*_KPC_ gene was identified using the CD-HIT v4.7 [34] data generated with cognac. The gene cluster containing coding sequences annotated by RAST v1.035 [31] as *bla*_KPC_ was identified. *bla*_KPC_ status was determined for each genome as having representation in the *bla*_KPC_ cluster.

## Supplemental table and figures

**Table S1:** Number of each facility type by state and how many facilities were represented in whole-genome sequencing (in parentheses).

| Facility type | CT | MN | TN |
| --- | --- | --- | --- |
| ACH | 30 (19) | 50 (16) | 92 (31) |
| LTACH | 3 (0) | 2 (1) | 10 (4) |
| SNF | 224 (0) | 366 (5) | 309 (0) |
| Other | 7 (0) | 96 (1) | 48 (1) |
| Unknown | 0 (2) | 0 (12) | 0 (4) |

**Figure S1:**
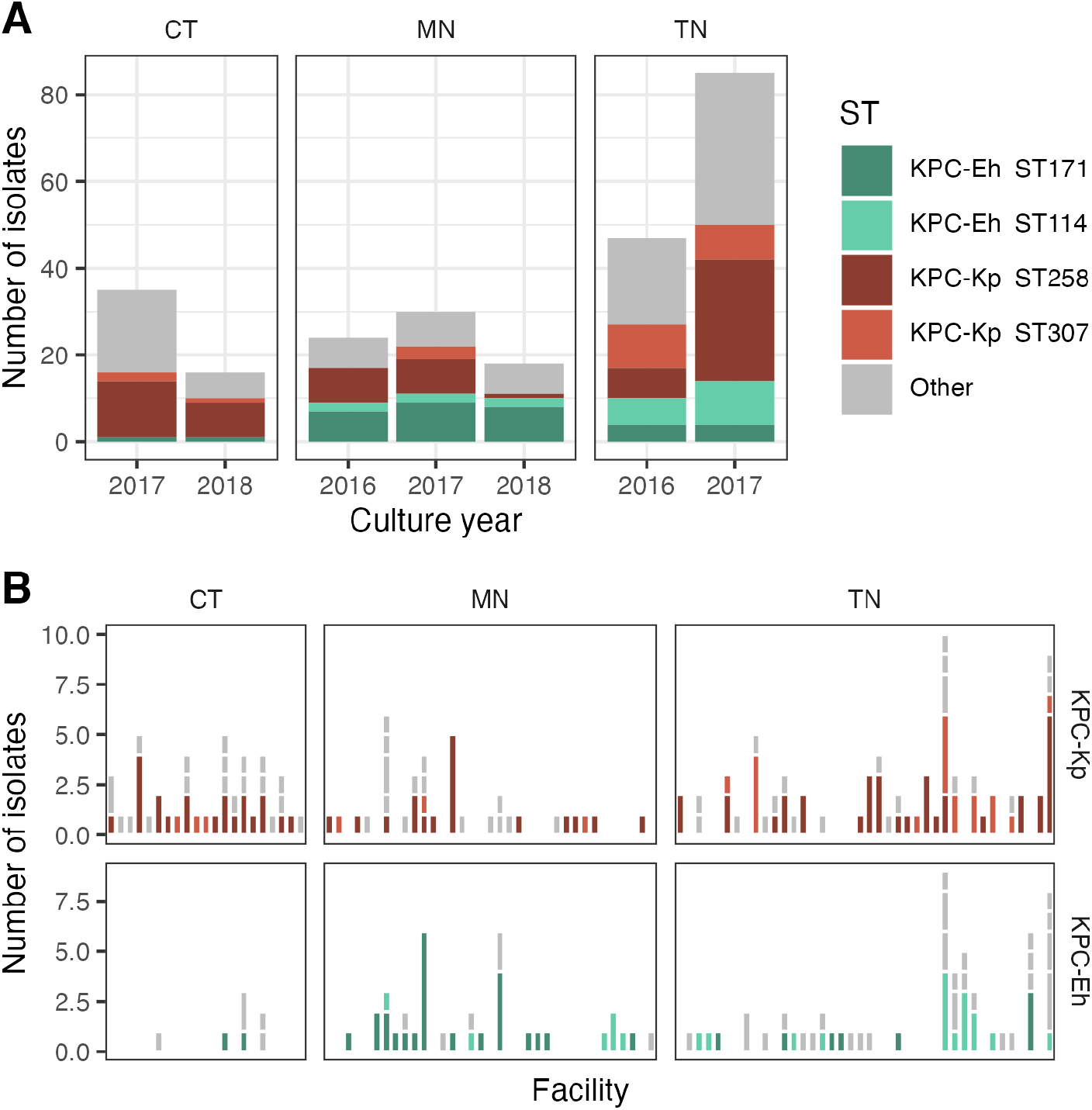
Distribution of species and sequence types of sequenced isolates from each state. (A) Across time. (B) Across healthcare facilities with at least one identified CRE with *bla*KPC. X axis shows facilities. Grey is any other ST of the species corresponding to that faceted plot. Notes: The MuGSI catchment area for MN is Hennepin and Ramsey counties. Four counties in the Memphis-Delta Emergency Medical Services (EMS) and 8 in Northeastern TN were excluded due to extensive healthcare utilization across state lines; the MuGSI catchment area is TN EMS Region 5, an 8-county region encompassing the metropolitan Nashville area, of TN. CT=Connecticut; MN=Minnesota; TN=Tennessee; ST=sequence type; Eh=*E. hormaechei*; Kp=*K. pneumoniae*.

**Figure S2:**
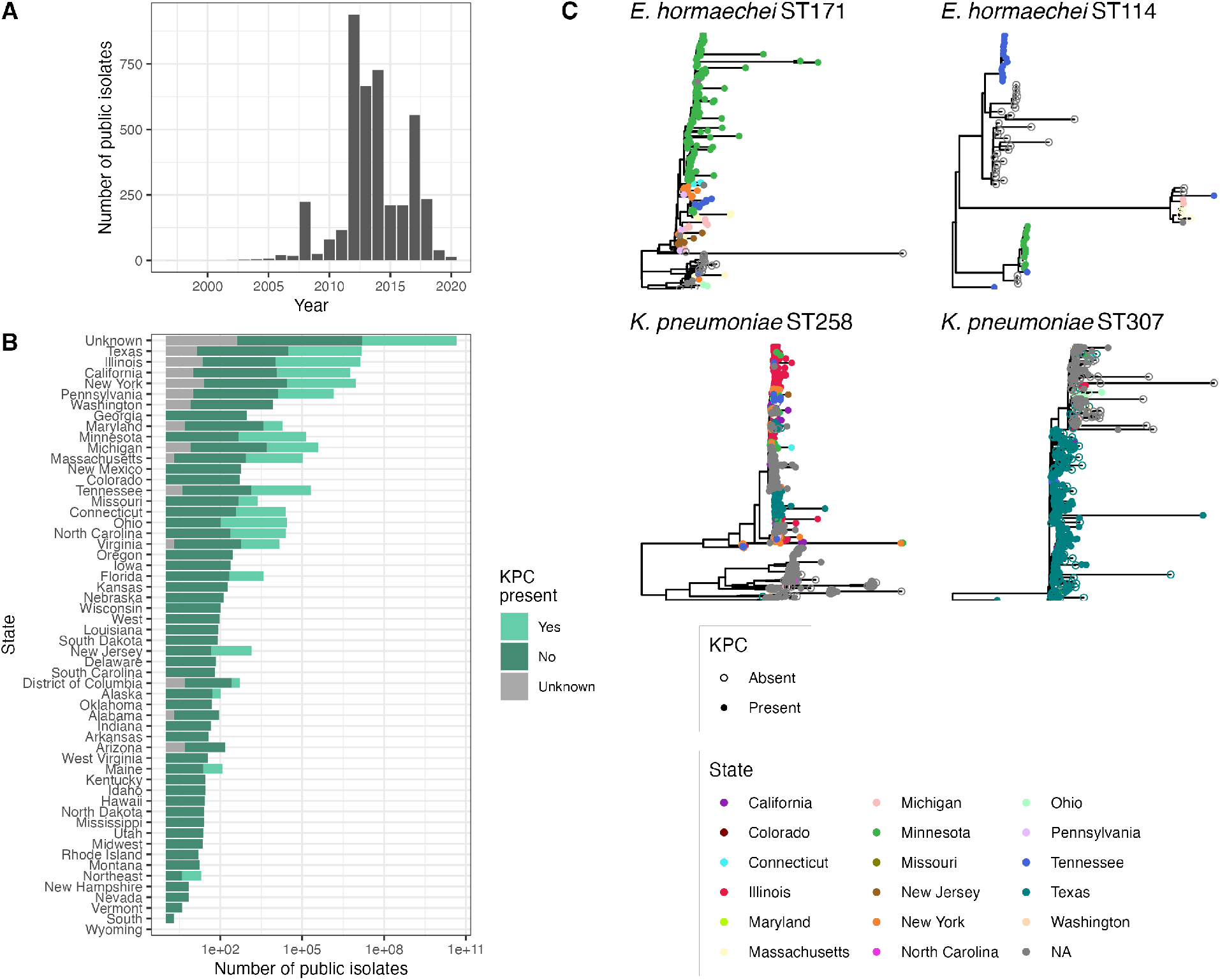
Information about public isolates downloaded from the PATRIC database that were included for context in the investigation of importation. (A) Distribution of year of isolation for isolates where this information was provided. (B) Distribution of location and KPC presence/absence. (C) Phylogenies used to identify importation events, including isolates from this study. ST=sequence type.

**Figure S3:**
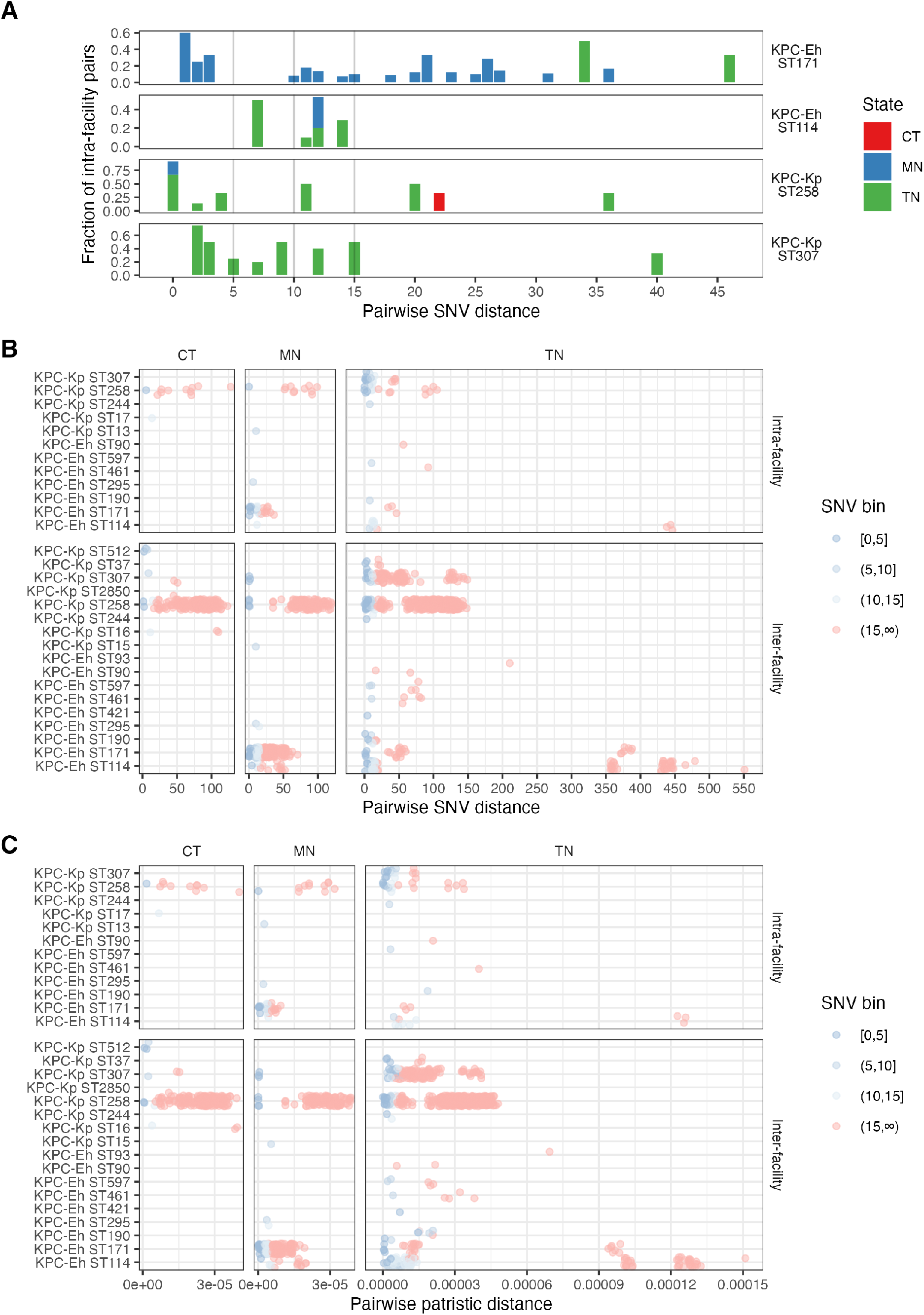
Inter-facility genomic analysis. (A) Fraction of intra-facility isolate pairs for pairwise single nucleotide variant (SNV) distances to help identify pairwise SNV distance cutoffs. We expect there to be an enrichment in closely related intra-facility pairs under the assumption that intra-facility transmission is more common than inter-facility transmission. This plot helps us identify potential pairwise SNV distance thresholds by looking for a SNV distance where there is a decrease in the fraction of intra-facility isolate pairs (around 5 and 10 appear to be reasonable thresholds for a sensitivity analysis). Vertical lines at 5, 10, and 15 SNVs represent potential cutoffs for determining what isolates are considered closely related; 3 thresholds were chosen as a sensitivity analysis. (B) Pairwise SNV distances of all STs. Within an ST, smaller pairwise SNV distances indicate potential local transmission, while larger pairwise SNV distances are not indicative of local transmission. Varying extents of transmission were captured for different states and STs. Putative local transmission events are those in the tail of the distribution (lower pairwise SNV distances). (C) Pairwise patristic distances are concordant with pairwise SNV distance thresholds. CT=Connecticut; MN=Minnesota; TN=Tennessee; ST=sequence type; Eh=*E. hormaechei*; Kp=*K. pneumoniae*; SNV = single nucleotide variant.

**Figure S4:**
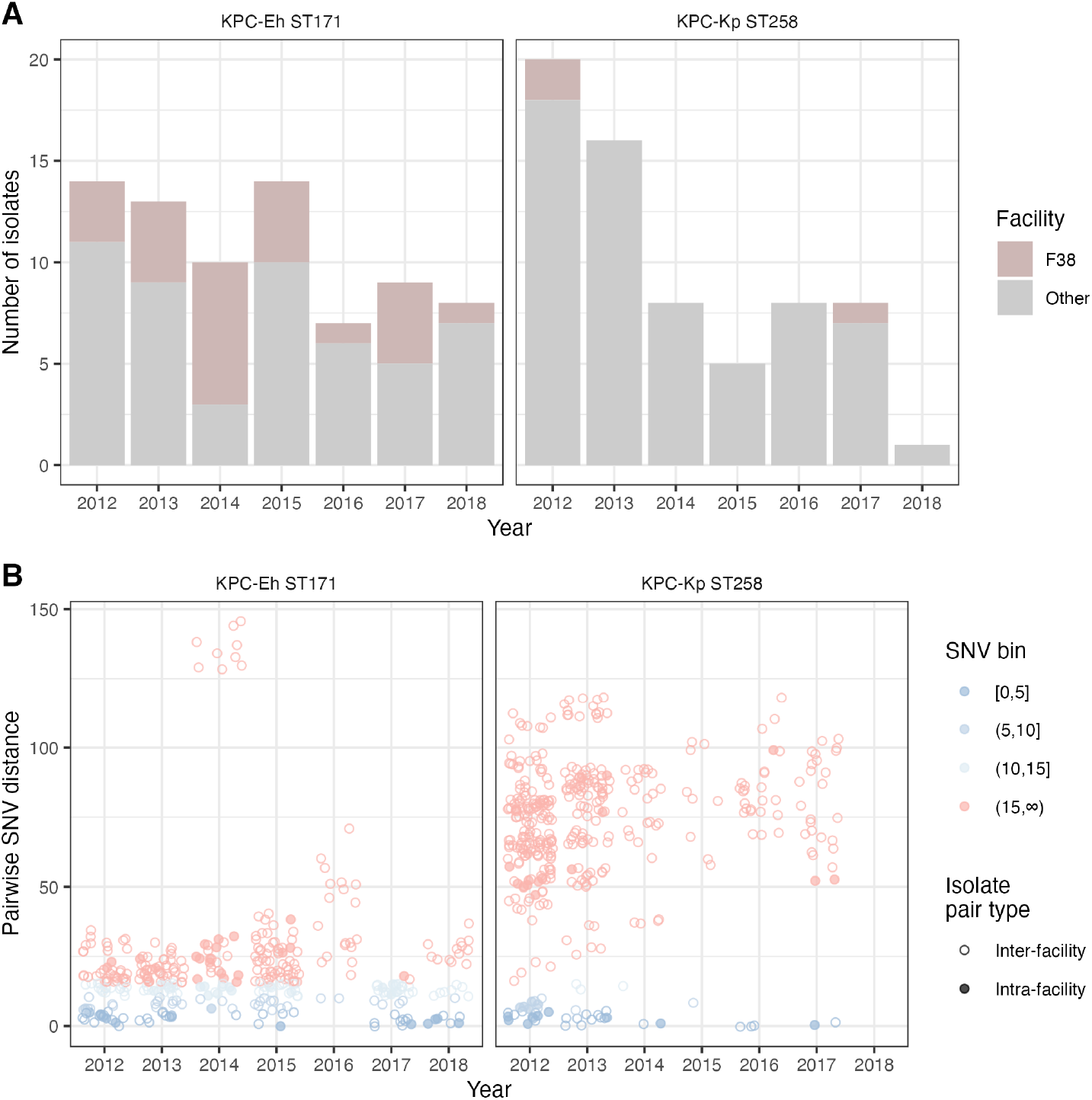
Minnesota (MN) isolates over time. (A) Number of ST171 and ST258 isolates over time. (B) Pairwise SNV distance of isolates over time. ST=sequence type; Eh=*E. hormaechei*; Kp=*K. pneumoniae*.

